# Diverticular disease genome-wide association meta-analysis identifies 257 novel loci, implicates structure and motility

**DOI:** 10.1101/2025.03.27.25324777

**Authors:** Christopher J. Neylan, Michael G. Levin, Gabrielle Shakt, Katherine Hartmann, Katherine Beigel, Sam Khodursky, Sarah Abramowitz, Emma E. Furth, Robert O. Heuckeroth, Scott M. Damrauer, Lillias H. Maguire

## Abstract

**Background and aims:** Diverticular disease is a common and morbid complex phenotype influenced by both genetic and environmental risk factors. The aim of the current study is to elucidate the genetic architecture of diverticular disease and to corroborate those results with tissue-based analysis.

**Methods:** We performed the largest genome-wide association study (GWAS) meta-analysis for diverticular disease. We employed multiple downstream analytic strategies, including tissue and pathway enrichment, statistical fine-mapping, protein quantitative trait loci, drug-target investigations, and linkage disequilibrium score regression to prioritize causal genes. We utilized single-cell RNA sequencing data and quantitative analysis of surgically resected colon specimens to corroborate our computational biology results.

**Results:** Our GWAS meta-analysis included 1.3 million individuals across three cohorts and identified 565 independent signals (257 novel) associated with diverticular disease. These were mapped to 432 unique genes. Several lines of evidence linked these genes to alterations in connective tissue biology and colonic motility. Analysis of single-cell RNA sequencing data revealed that prioritized diverticular disease-associated genes are enriched for expression in colonic smooth muscle, fibroblasts, and interstitial cells of Cajal. Quantitative analysis of surgically resected colon specimens found a substantial (43%) reduction in the density of elastin present in the sigmoid colon among those with severe diverticulitis relative to those with diverticulosis.

**Conclusion:** Diverticular disease is predominantly a disorder of connective tissue biology and colonic motility. Specifically, decreased density of elastin in the muscularis propria may predispose not to diverticulosis but to diverticulitis itself.

## Introduction

Diverticulitis is commonly understood as an inflammatory phenomenon that is triggered by an unknown inciting event in a patient with diverticulosis.^1,2^ Certain environmental risk factors (such as the Western diet) have well-established associations with diverticulitis, but disentangling causal risk factors for diverticulosis from those for diverticulitis has been difficult.^3–8^ More recently, a genetic etiology of diverticular disease was established, with twin studies estimating 40-53% heritability.^9,10^ Genome-wide association studies (GWASs) implicated 150 genetic risk loci to date. Polygenic risk scores derived from these GWASs correlate with diverticulitis incidence and severity, however these studies are limited by relatively homogenous genetic populations and persistent inability to distinguish diverticulosis from diverticulitis.^9–15^

In the current study, we perform a GWAS meta-analysis for diverticular disease (DD) and identify 257 loci that were not previously described for DD. In subsequent analyses we prioritize genes, pathways, and proteins that may be causally involved in the development of diverticulitis. We identify proteins that can serve as targets for drug repurposing and we integrate single cell RNA sequencing (scRNAseq) data to localize gene expression to individual cell types. These analyses implicate abnormal extracellular matrix composition (particularly elastin and related molecules) and disordered smooth muscle motility in the pathogenesis of diverticulitis. Building on this finding, we use linkage disequilibrium score regression to assess shared genetic liability with other traits, finding overlap with connective tissue diseases (such as hernia, pelvic organ prolapse, and abdominal aortic aneurysm), and the lack of overlap with intestinal inflammatory conditions such as ulcerative colitis and Crohn’s disease.

To investigate the hypothesis that extracellular matrix changes are involved in the development of diverticulitis *per se* rather than merely diverticulosis, we analyzed the elastin content in the colonic wall of surgical specimens obtained from individuals without diverticular disease, with asymptomatic diverticulosis, and with varying degrees of severity of diverticulitis. We find that individuals requiring emergency surgery for diverticulitis had significantly decreased elastin density throughout the entirety of the evaluated colon relative to all other groups, further supporting the theory of diverticulitis as a disease driven by connective tissue changes.

## Methods

### GWAS meta-analysis

To identify genetic variants associated with diverticular disease, we performed a meta-analysis of three publicly available genome-wide association studies (GWASs) conducted in separate discovery cohorts (the Million Veteran Program [MVP], the United Kingdom Biobank [UKB], and Finngen) (Extended Data Fig. 1).^16–18^ Details of the Department of Veterans Affairs (VA) MVP longitudinal health cohort, including its harmonized ancestry and race/ethnicity (HARE) approach, can be found elsewhere,^16,19^ as can descriptions of the Pan-UKB initiative^20,17,21^ and FinnGen.^18^ In each source GWAS, diverticular disease was identified by Phecode 562, a parent code for a group of nine International Classification of Disease (ICD)-10 codes and 19 ICD-9 codes specific to diverticulosis and diverticulitis.^22^

Quality control was performed on each set of summary statistics to remove variants with poor imputation scores (*r*^2^ < 0.3 in MVP; *r*^2^ < 0.8 in UKB and FinnGen, according to thresholds set in the original GWASs) or erroneous p-values or allele frequencies (e.g. > 1 or < 0). Meta-analysis was performed with a fixed-effects, inverse-variance weighted model using METAL.^23^ Meta-analyzed data was then quality-controlled and cleaned using GWASinspector (Supplementary Data Fig. 1).^24^ SNPs with a minor allele frequency (MAF) < 0.01 were discarded, strand flips were corrected, and variants were removed if they were missing crucial values, duplicated, or ambiguous.

### Definition of loci and secondary signals

From the meta-analyzed overall cohort, loci were defined around genome-wide significant variants (p < 5 x 10^−8^) via local clumping with a 500kb window. A single lead variant (or index variant) was retained per locus (Extended Data Table 1). To identify additional, independently significant variants within a locus (secondary signals), we performed statistical fine-mapping using CARMA, using the 1000 Genomes Project Phase 3 European LD reference panel.^25^ A fine-mapped variant was considered a secondary signal if it fell within a defined credible set and had the highest PIP within its credible set. Index variants and secondary signals were carried forward for downstream analysis, however if an index variant was not the highest PIP SNP in its credible set, the index variant was removed and only the secondary signal from that credible set was carried forward for analysis.

### Gene mapping and prioritization

An ensemble approach considering several methods was used to map each signal to a gene. First, the nearest gene to the signal was identified using *gwasRtools* package in R.^26^ Next, MAGMA was run using the *magmaR* package to identify a gene associated with the signal based on the 1000 Genomes Project Phase 3 EUR imputation panel.^27^ Third, OpenTargets annotations were analyzed to determine whether the signal lies in a known coding region or regulatory region for a gene.^28^ Finally, multi-trait colocalization was performed using HyPrColoc to identify colocalization between the signals and eQTLs in the eQTLGen Phase I cis-eQTL blood data.^29^ All genes mapped to each signal by these methods were then tabulated and a single gene was prioritized for each signal based on a process that prioritized coding variants over non-coding variants, and among non-coding variants gave preference to genes identified by multiple methods at the same signal. (Extended Data Table 2)

### Enrichment analysis

The list of genes to which signals were mapped was then subjected to a suite of enrichment analyses to gain insight into the biological function of the genes. First, utilizing Enrichr, the list of genes was analyzed for enrichment against numerous gene set libraries (e.g. the Gene Ontology Cellular Component dataset) (Fig. 1a, b).^30–32^ Then, FUMA (*GENE2FUNC* feature) was used to analyze tissue expression with the following parameters: all background genes, GTEx v8 tissue types (excluding MHC region), FDR multiple test correction (adjusted p < 0.05) (Fig. 1c).^33^

**Fig. 1.**
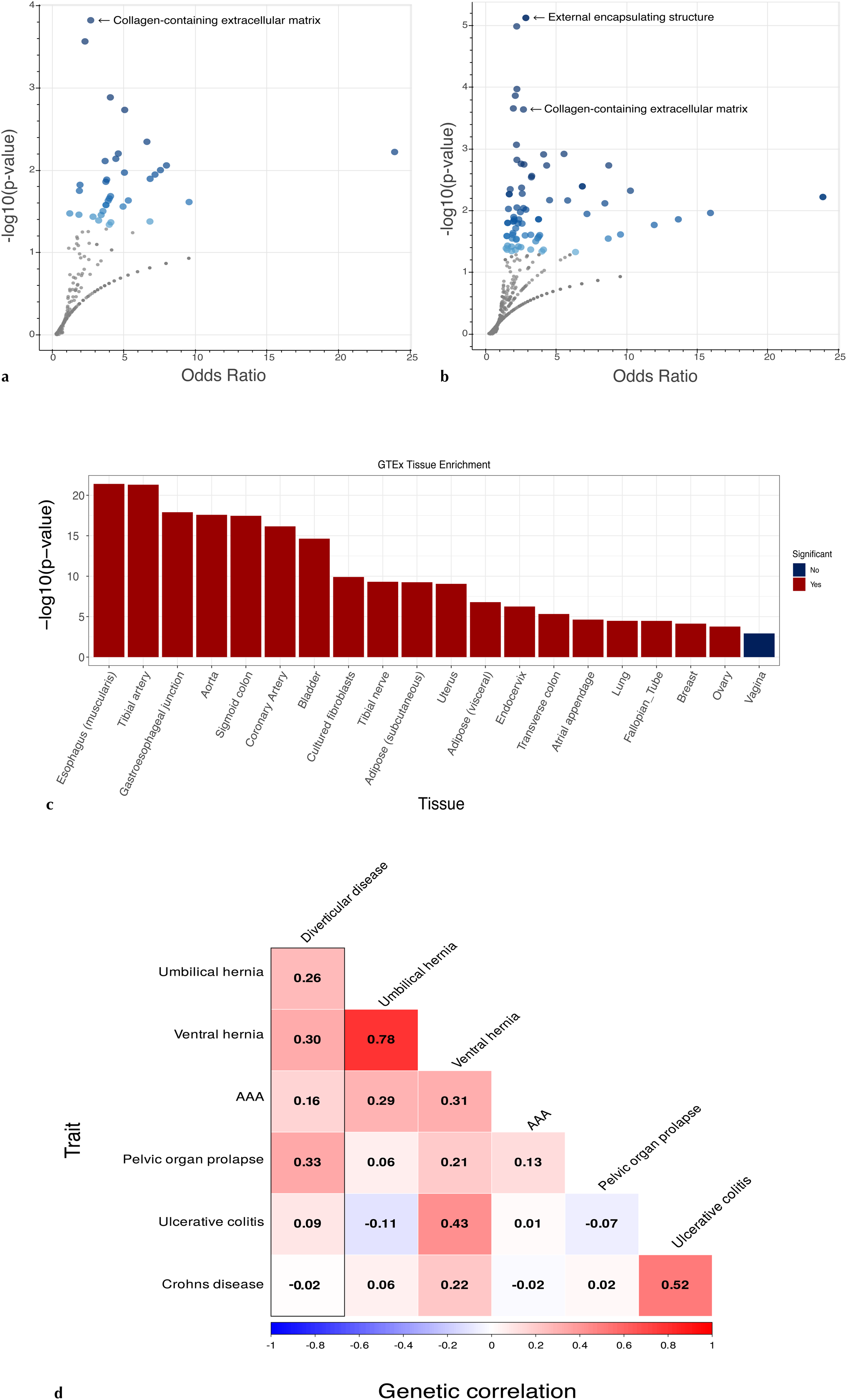
Biological implications of genetic variation associated with diverticular disease, showing enrichment of prioritized genes in **a,** GO Cellular Component (2025), **b,** COMPARTMENTS (2025), and **c,** GTEx (v8) tissues, followed by **d,** genetic correlation between diverticular disease and other phenotypic traits as assessed by linkage disequlibrium score regression. In volcano plots (a and b), each dot represents a term from the target gene set in which the prioritized genes were enriched; blue dots are significant (p-value < 0.05). In c, significance is determined by adjusted p-value < 0.05.

### Linkage Disequilibrium Score Regression (LDSC)

LDSC was used to estimate the heritability of diverticular disease and genetic correlation with select phenotypes.^24^ LD scores derived from the 1000 Genomes Project European samples were utilized. SNP heritability was transformed to the liability scale using a sample prevalence of 12.3% (the calculated prevalence in our data) and a population prevalence of 30% (the approximated epidemiologic prevalence of disease).^34^ Publicly available summary statistics were used for phenotypes other than diverticular disease (Supplementary Data Table 1).

### Protein drug targets

We performed a proteome-wide plasma Mendelian randomization screen to identify proteins with putatively causal effects on the development of diverticular disease, which could serve as targets for the repurposing of existing drugs. First, we identified genetic variants associated with plasma protein levels (protein quantitative trait loci, or pQTLs) in two large-scale plasma-proteomics studies (deCODE and UK Biobank Pharma Proteomics Project, hereafter UKB-PPP).^35,36^ Then we performed Mendelian randomization using pQTLs as instrumental variables and our diverticular disease GWAS meta-analysis summary statistics to instrument the outcome of diverticular disease. Instrumental variables were selected if they met the following criteria: genome-wide significance (p < 5 x 10^−8^), maximum linkage disequilibrium *r*^2^ of 0.01, and *cis* variant within a 500kb window of the respective gene. Wald ratios were used for instruments comprised of a single SNP, and inverse-variance weighted MR was used otherwise. Proteins were considered significant if the adjusted p-value was < 0.05, according to the Benjamini-Hochberg correction to maintain FDR < 0.05. Using a previously-generated list of 1,263 actionable targets, defined as proteins targeted by approved drugs or drugs in the clinical phase of development, we then identified actionable targets within our own results.^37^ To understand the biological implication of the significant proteins, we performed enrichment analysis of the identified proteins in the Reactome (2024) pathway database.

### Immunohistochemistry

Formalin-fixed, paraffin-embedded tissue blocks from surgically resected sigmoid colons were obtained from the Pathology Department of the Hospital of the University of Pennsylvania. The indication for surgery was stored in a variable termed ‘Patient Classification’ which took the values of ‘No disease’ for an incidental resection (e.g. iatrogenic sigmoid injury) in which the colon did not have diverticular disease; ‘Diverticulosis’ for an incidental resection in which the patient had diverticulosis but not diverticulitis; ‘Emergent diverticulitis’ for free colonic perforation; or ‘Non-emergent diverticulitis’ for any acutely symptomatic diverticulitis, recurrent diverticulitis asymptomatic at the time of surgery, or colonic fistulization. Patients with known connective tissue disorders or confounding colonic pathology (e.g. inflammatory bowel disease) were excluded. Microscopy slides were prepared from tissue samples taken from areas of grossly apparent disease and from the margins of the surgical specimen (the area furthest from gross disease). Stains for elastin (Verhoeff’s stain), the smooth muscle myosin heavy chain 11 (MYH11), and collagen (Sirius Red) were applied and the density of elastin and collagen present in the tissue was quantified with the aid of pixel classifiers (Supplementary Data Fig. 2).^38^

### Single cell RNAseq analysis

Cell-level gene expression was ascertained from single cell RNA-sequencing (RNAseq) of the human colon in two previously published datasets.^39,40^ In the Hickey et al. (2023) dataset, we restricted our analysis to samples from the sigmoid colon and re-annotated “Myofibroblasts/SM 1,” “Myofibroblasts/SM 2,” and “Myofibroblasts/SM 3” as ‘Myofibroblasts’. We employed the single-cell disease-relevance score (scDRS) to evaluate expression of our GWAS-associated genes in individual cells.^41^ We then applied k-means clustering and generated clustered dot plots using the R package scCustomize in order to assess patterns of gene expression.^42^ We visually identified clusters of genes with enriched expression in any cell type noted to be significant in scDRS, then generated heatmaps of gene expression for all genes that comprised the relevant clusters, then performed pathway analysis of the genes the comprised the relevant clusters using Enrichr.

## Results

### GWAS meta-analysis identified 394 loci

Our meta-analysis consisted of 1,371,893 individuals in the combined MVP, UKB, and FinnGen cohorts, of whom 157, 944 (12%) had diverticular disease and 1,213,949 (88%) did not. In the overall cohort, 1,201,833 (87%) individuals were genetically similar to the European reference population (EUR); 114,236 (8.3%) to the African reference population (AFR); 46,867 (3.4%) to the admixed American population (AMR); 7,587 (0.55%) to the Central/South Asian (CSA) population; and 1,370 (0.01%) to the Middle Eastern (MID) population. We obtained single-variant association statistics for a total of 51,738,151 SNPs. After removing variants that did not pass quality control (see Supplementary Data Fig. 1) or had a minor allele frequency (MAF) < 0.01, we carried forward 19,070,016 variants for analysis. We identified 394 loci defined around index variants that were genome-wide significant (p < 5 x 10^−8^) for their association with diverticular disease, of which 257 (65%) were novel (Extended Data Table 1). After performing statistical fine-mapping with CARMA to identify secondary signals, a total of 565 independent signals were carried forward for analysis (Extended Data Table 2).

### Gene mapping and biological function

The 565 independent signals were mapped to 432 unique genes. While many of these genes were mapped based on distance to the specified signal (and thus potential nearby regulatory function), a smaller subset achieved a higher threshold of established roles in human diseases: 14 (3%) were missense variants and 41 (9%) were coding or well-annotated regulatory variants (Extended Data Table 2). Interrogation of the Gene Ontology Cellular Component (2025) pathway database showed the prioritized genes were significantly enriched in extracellular matrix formation (Fig. 1a), a result which was corroborated by analysis with other pathway databases (Fig. 1b). Specific mapped genes involved in extracellular matrix remodeling include *ELN*, *LTBP1*, *ECM1*, *LOXL1, COL* family and *ADAM* family genes. Other mapped genes were involved in smooth muscle or interstitial cells of Cajal function, such as *ANO1, RYR2,* and *VIPR1,* thereby implicating both colonic structure and motility. Analysis of the DD-associated genes across human tissue types in GTEx (v8) revealed significant enrichment in smooth muscle-containing organs such as the sigmoid colon, transverse colon, esophagus, bladder, uterus, and arteries (Fig. 1c).

### Linkage Disequilibrium Score Regression

The unadjusted, observed heritability (*h*^2^) of diverticular disease was 0.09 (standard error = 0.005, p = 3.2 e −59) with a liability-scale SNP heritability (*h^2^_SNP_*) of 0.32. Moderate genetic correlation was observed between diverticular disease and connective tissue phenotypes such as hernia and pelvic organ prolapse, while weak genetic correlation was observed with abdominal aortic aneurysm (AAA), and negligible correlation with ulcerative colitis and Crohn’s disease (Fig. 1).

### Protein drug targets

Proteome-wide Mendelian randomization analysis, using instruments derived from pQTLs identified in multiple data sets, to link diverticular disease GWAS associations to serum protein levels identified a total of 188 proteins whose predicted levels were associated with genetic liability to diverticular disease (Fig. 2; Supplementary Data Tables 2 and 3). Many of these proteins whose abundance in plasma was associated with diverticular disease-associated SNPs directly overlapped with our independently mapped genes in GWAS analysis, including *ELN, COL15A1*, and *COL6A1*, while other proteins corresponded to the same sub-families as mapped genes (e.g. *LTBP4* and *LOXL3* were identified by the Mendelian randomization analysis, corresponding to *LTBP1* and *LOXL1* identified in the GWAS analysis). Analysis of the 188 proteins using the Reactome 2024 database (utilizing Enrichr) revealed enrichment in the extracellular matrix proteins and elastin-forming processes (Fig 2c), mirroring the enrichment analysis for our mapped genes. It should be noted that this plasma MR experiment identifies circulating proteins, not proteins measured in colonic tissue. However, extracellular matrix proteins are often released into circulation during disease states, making a segment of colon with diverticulitis a plausible source of the observed proteins.^43^

**Fig. 2.**
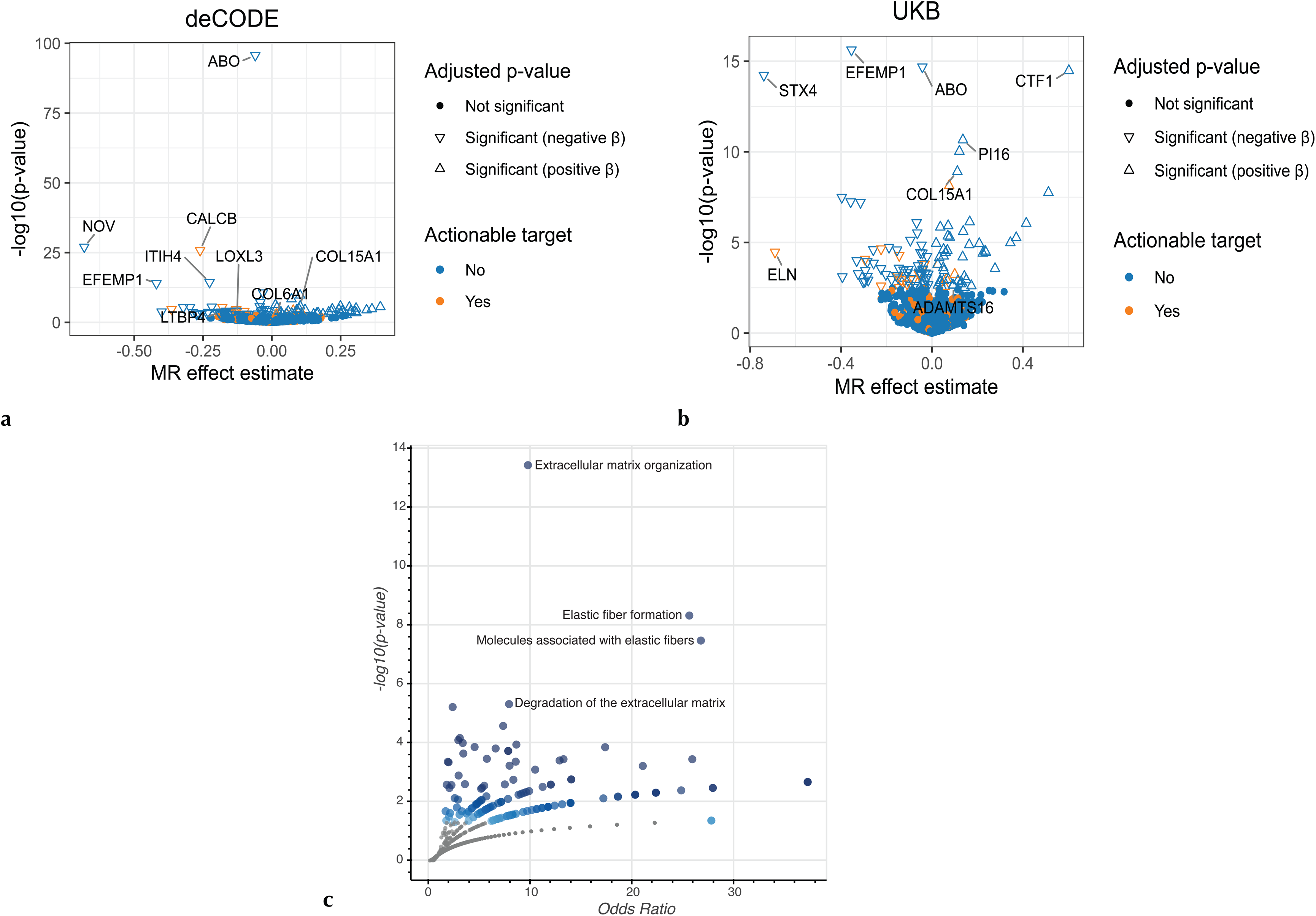
pQTL Mendelian randomization identifies 188 proteins associated with diverticular disease in the **a,** deCODE and **b,** UKB-PPP databases. MR effect estimate is the natural logarithm of the odds ratio. Positive beta means increase serum protein abundance is positively associated with diverticular disease. **c**, shows the biological processes associated with the 188 proteins in the Reactome (2024) database. Blue points have a p value < 0.05

Of the 188 proteins found to associate with diverticular disease, 25 (13.3%) were known drug targets, including *ELN* and *TNF* products (Fig. 2). These provide the possibility of actionable targets, allowing the repurposing of existing drugs against these gene products.

### Immunohistochemistry

To further define anatomic findings in diverticular disease, assess the plausibility of links to elastin, and appreciate differences between individuals with diverticulitis relative to individuals with diverticulosis, we analyzed colon surgical resections from 24 patients (78 microscopy slides). Of the 24 individuals, 13 (54%) has diverticulitis, 5 (21%) had diverticulosis without diverticulitis, and 6 (25%) were without diverticular disease. Among the 13 individuals with diverticulitis, 3 (23%) had life-threatening diverticulitis requiring emergency surgery, while 10 required elective, non-emergent surgery. A representative *MYH11* antibody-stained slide from a non-emergency diverticulitis patient demonstrates severe attenuation of the muscularis propria overlying the diverticulum (Fig. 3). Note that the figure does not depict the diverticulitis-causing (i.e. perforated) diverticulum, but rather an asymptomatic diverticulum in the resected region. Because the observed diverticulum is not at the exact site of perforation, it is less likely that attenuated muscularis propria is secondary to the perforation (or its associated inflammation), and more likely that it was present in the tissue prior to the perforation, raising the possibility that this muscular change is present in many regions of the sigmoid colon of the sampled patient.

**Fig. 3.**
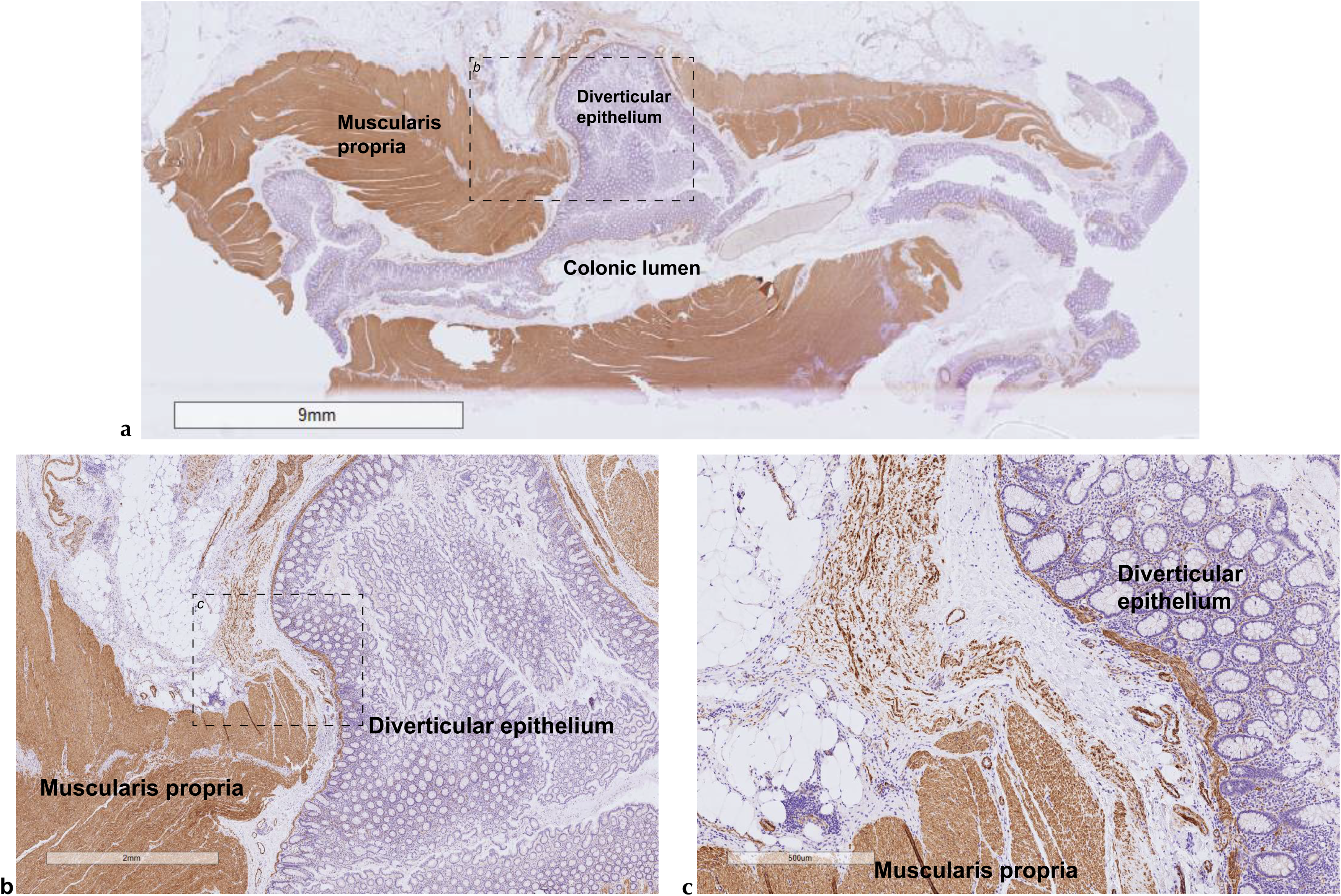
MYH11 antibody staining. (brown) of colonic tissue resected during surgery from a patient with diverticular disease, demonstrating thinning of the muscularis propria overlying the diverticulum at **a,** 1x magnification, **b,** 5x magnification, and **c,** 20x magnification. The area outlined with dashed lines in panel (a) shows the borders of panel b, and the area outlined with dashed lines in panel b shows the borders of panel c. Hematoxylin (purple) was used as a counter-stain.

To test the hypothesis that weakness of the elastin extracellular matrix could predispose to diverticulitis, as suggested by our GWAS results, we analyzed elastin fibers in the muscularis propria of resected human sigmoid colons. Several areas were sampled for analysis (Extended Data Fig. 2). In diverticulitis patients, samples were taken from: around the perforated diverticulum, and separately from around a diverticulum at the surgical margin (i.e. the region furthest from the point of diverticulitis). In diverticulosis patients, samples were taken from: around a diverticulum (i.e. areas of gross disease), and separately from grossly healthy areas (i.e. areas without diverticulosis). In patients without diverticular disease, samples were taken from areas that appeared grossly healthy. Across all samples, the density of elastin fibers in the muscularis propria was markedly decreased among individuals who required emergency surgery for diverticulitis compared to all other groups (Fig. 4a). In attempt to exclude the local effects of diverticulitis-associated inflammation and uncover potentially genetically-mediated differences between individuals with and without diverticulitis, we compared the density of elastin present at the surgical margin in patients with emergency diverticulitis (location 5 in Extended Data Fig. 2) to a) the density of elastin present immediately adjacent to diverticula in individuals with diverticulosis (location 2 in Extended Data Fig. 2), and b) the density of elastin present in individuals without diverticular disease (location 1 in Extended Data Fig. 2). These are the appropriate anatomic sites for comparison because they create the most stringent contrast: the site most likely to have normal elastin density in individuals with diverticulitis (the site furthest from the perforation) compared to the site least likely to have normal elastin density in individuals with diverticulosis (the site immediately adjacent to the diverticulum). To perform these comparisons, we constructed a multivariable linear regression (controlling for age and sex), which revealed that those requiring emergency surgery for diverticulitis had a 41% reduction (95% CI −66% to −17%, p = 0.001; Fig. 4b) in elastin density in the muscularis propria (sampled at location 5, away from the site of diverticulitis) relative to patients without diverticular disease (sampled at location 1), and a 43% reduction (95% CI −69% to −17%, p = 0.002) in elastin density compared to those with diverticulosis (sampled at location 2) (Fig. 4). Our analyses examining collagen content reveal no significant differences between groups (Extended Data Fig. 3).

**Fig. 4.**
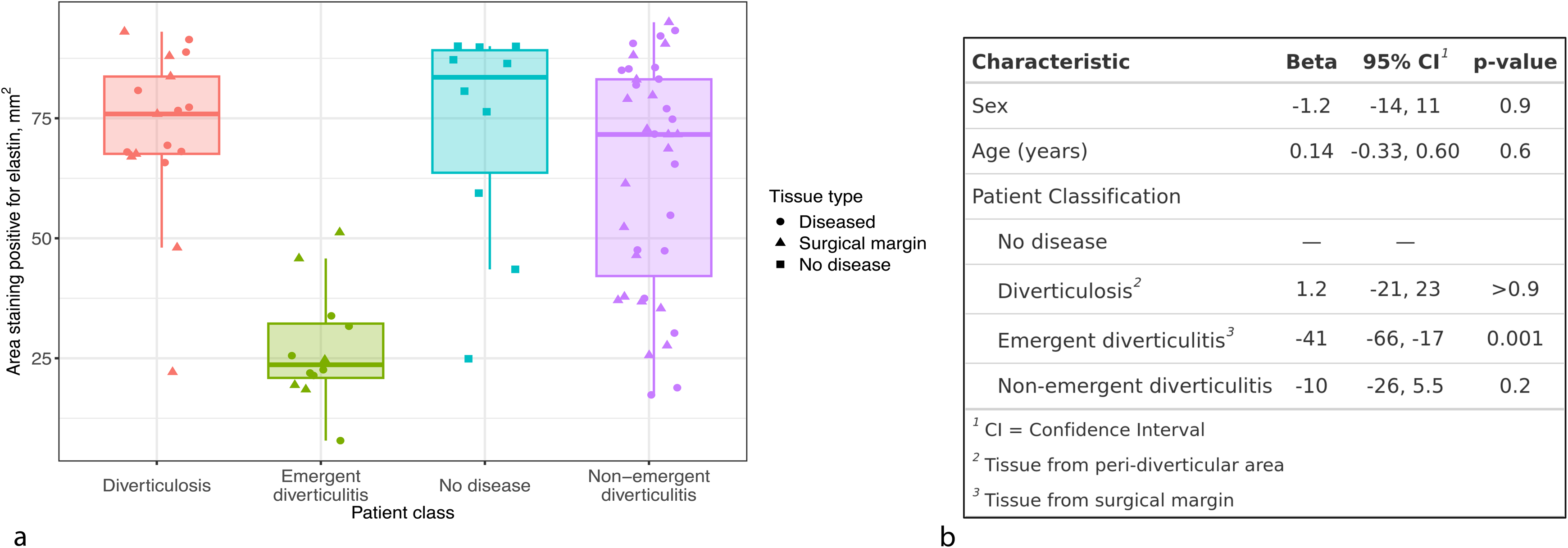
Elastin quantification. **a,** Each dot represents the measured elastin density on a prepared slide, with boxplot showing median elastin density (horizontal line) by patient class, interquartile range (IQR; the borders of the box) and whiskers extending 1.5*IQR from the end of the box. **b,** Multivariable linear regression showing a 41% reduction in elastin density among patients undergoing emergency diverticulitis surgery compared to those without disease. Peri-diverticular area corresponds to location 2 in Extended Data Fig.6; surgical margin corresponds to location 5 in Extended Data Fig.6.

### Single cell RNAseq analysis

In order to understand which biological processes may be disrupted by the genetic variation we found in our GWAS, we identified the specific cell types in which diverticular disease-associated genes are expressed under normal circumstances. We accomplished this by analyzing two single cell RNAseq datasets (Hickey et al. and Wright et al.) obtained from patients without known diverticular disease. We found that, when looking specifically at expression patterns in the sigmoid colon, DD-associated genes are abundantly expressed in fibroblasts, smooth muscle cells, interstitial cells of Cajal (ICC), neurons (Fig. 5).

**Fig. 5.**
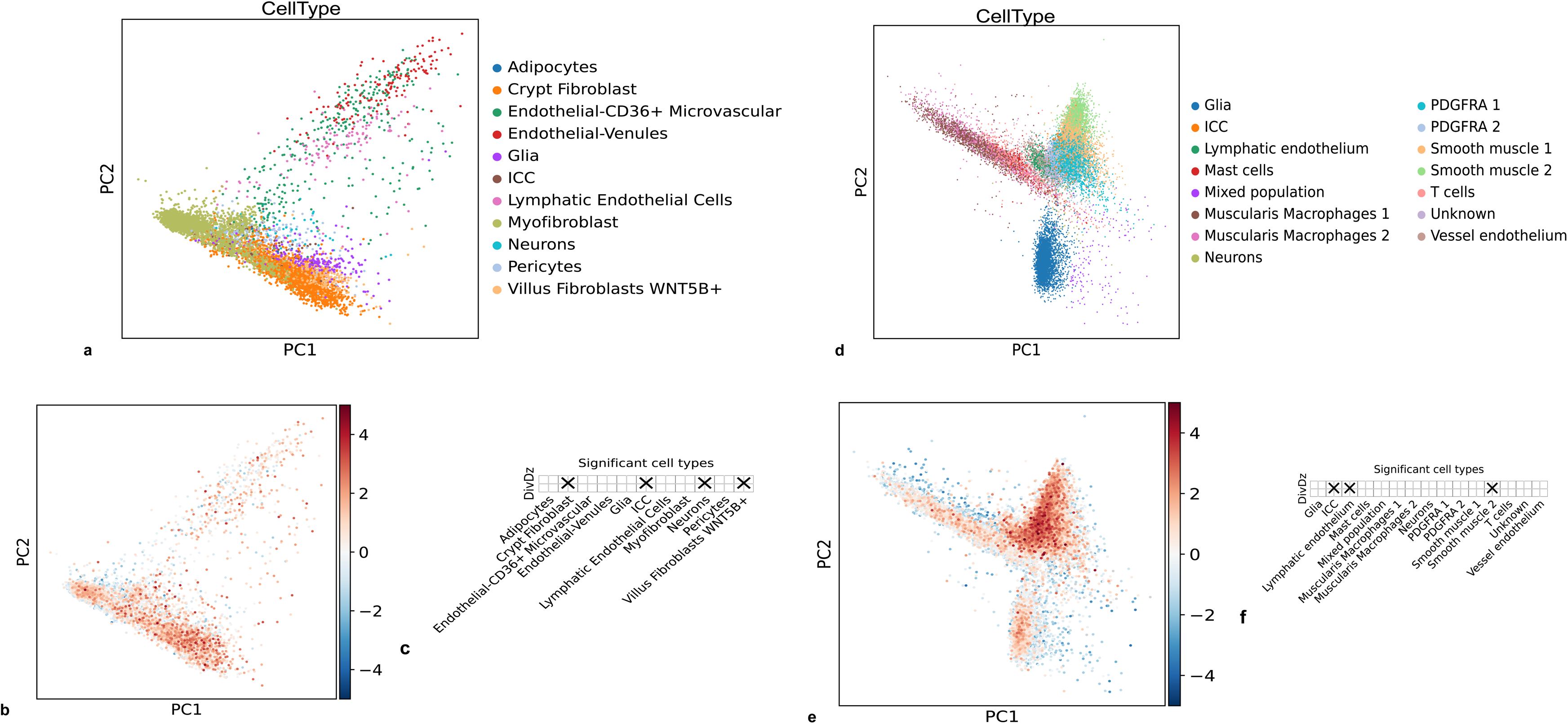
Cell-level localization of diverticular disease-associated genes. obtained with scDRS in the Hickey et al. (**a,b,c**) and Wright et al. (**d,e,f**) single-cell RNAseq datasets showing **a,d,** Dimensional reduction of analyzed cells, **b,e,** Relative cell-level expression of diverticular-disease associated genes, **c,f,** Cell type-gene expression associations, with X denoting significantly (FDR < 0.001) increased expression of diverticular disease (DivDz)-associated genes in the indicated cell type.

We then sought to determine which among the 432 DD-associated genes are responsible for the enriched expression in these cell types. In order to do this, we constructed dot plots with k-means clustering (Fig. 6) to narrow the 432 genes into differentially-expressed clusters, and further analyzed the highly-expressed clusters with heatmaps showing gene-level expression patterns (Fig. 7). This revealed high expression patterns of *ANO 1,* a calcium activated chloride channel that plays a role in pacemaker activity of the intestine, in the interstitial cells of Cajal.^44^ It also revealed enriched expression *CACNB2* and *RYR2,* both of which are calcium channels involved in muscle contraction, in colonic smooth muscle ^32,45^

**Fig. 6.**
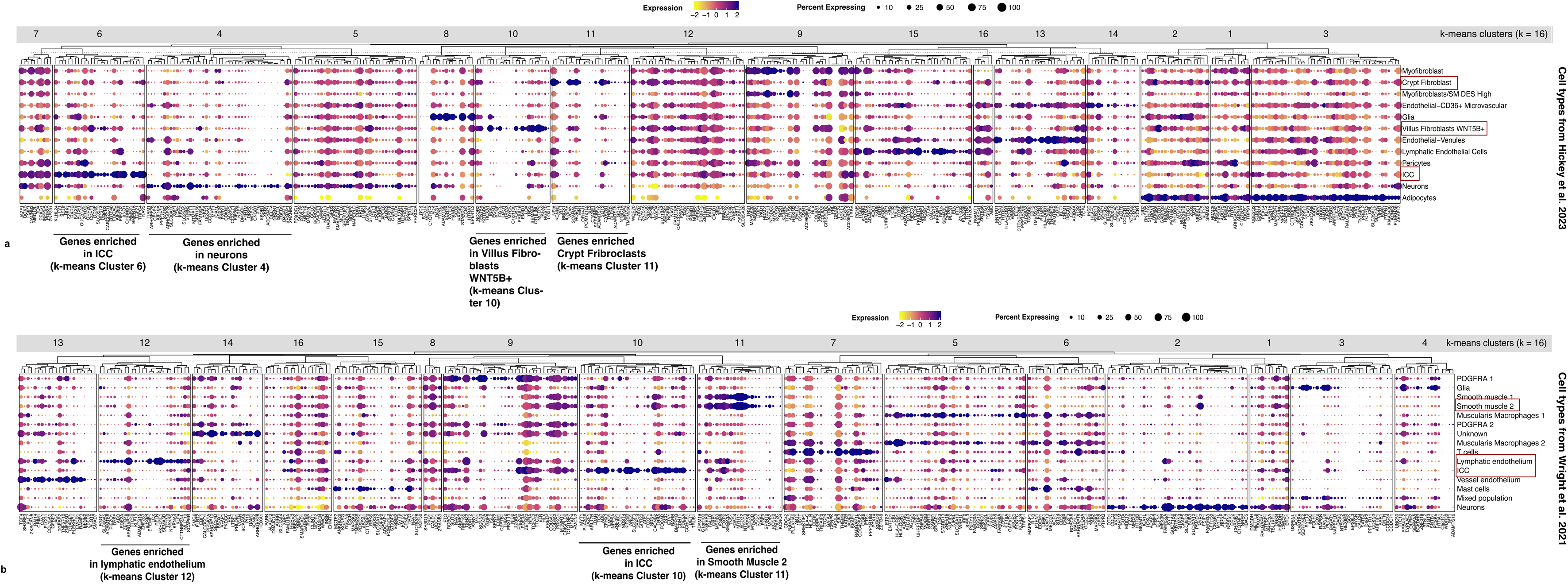
Dot plots showing expression patterns of diverticular disease-associated genes with k-means clustering applied, in the **a,** Hickey et al. and **b,** Wright et al. scRNAseq datasets. Mean expression for each gene across all samples is set to zero, and scaled expression is shown as standard deviations above or below the mean.

**Fig. 7.**
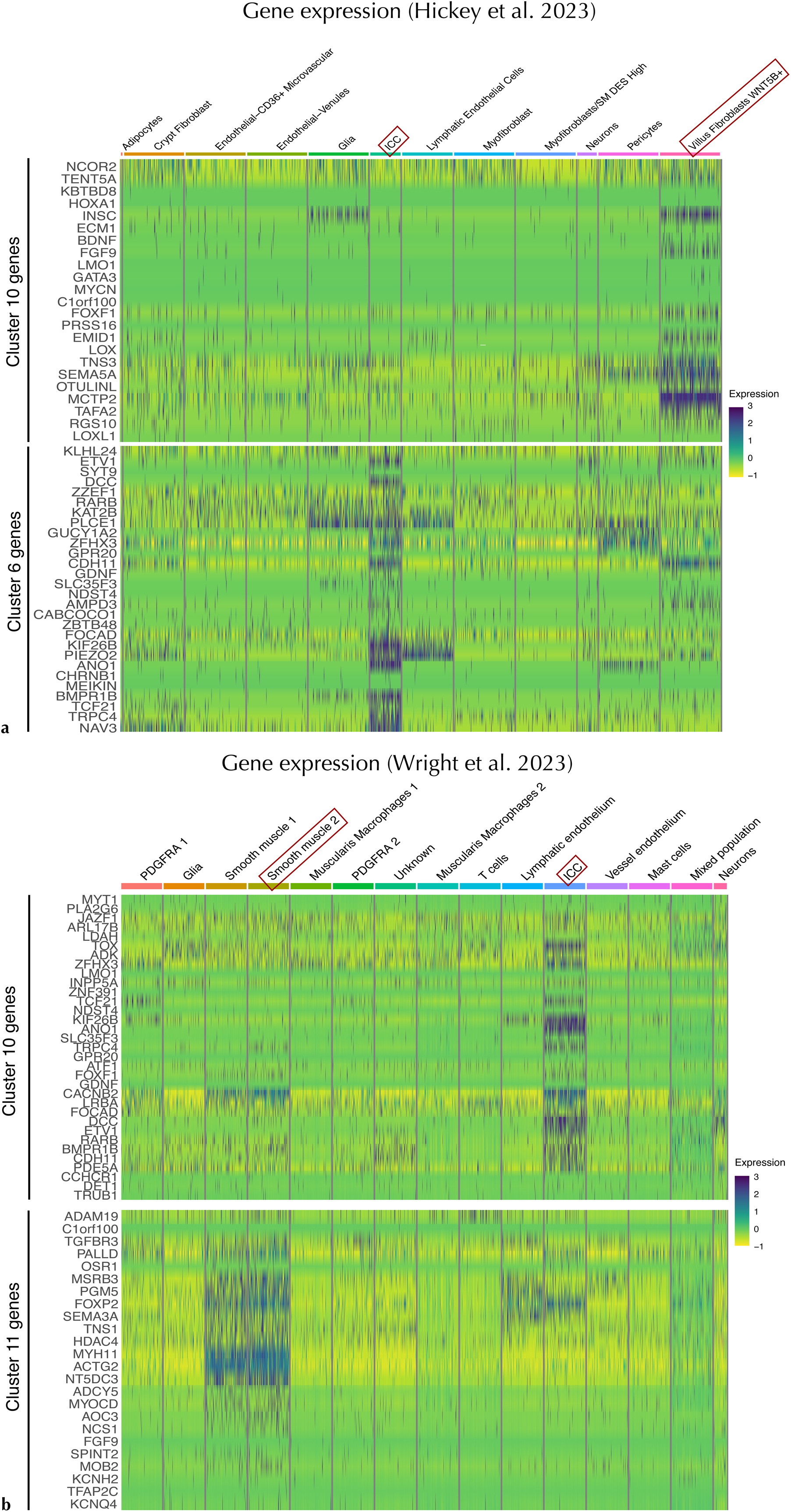
Heatmaps showing cell-level expression of select gene clusters by cell type in **a,** Hickey et al. WNT5B+ fibroblasts (cluster 10) and ICC (cluster 6) and **b,** Wright et al. ICC (cluster 10) and smooth muscle cells (cluster 11). Boxes denote cell types in which gene expression was enriched (see Fig. 6). Mean expression for each gene across all samples is set to zero, and scaled expression is shown as standard deviations above or below the mean.

In addition to gene-level expression using heatmaps, we analyzed all genes in highly-expressed clusters via pathway analysis (Extended Data Fig. 4), which implicated the biological processes of nerve conduction and extracellular matrix formation, all together suggesting that the diverticular-disease associated genes identified by our analysis have roles in maintining normal colonic motility and structure (these proposed biological mechanisms are illustrated in Extended Data Fig. 5.)

## Discussion

The current study offers multiple lines of evidence that diverticular disease (DD) is predominantly a disorder of connective tissue biology and colonic motility, in contrast to historical hypotheses that DD is an infectious or inflammatory disease. This assertion is concordant with more recent genomic work and with longstanding clinical associations between DD and connective tissue disorders, both rare (e.g. Ehlers-Danlos and Williams syndromes) and common (e.g. hernia and pelvic organ prolapse).^12,46–48^ We began with a GWAS meta-analysis for diverticular disease (DD) that identified 257 novel loci, more than doubling the number reported in the literature. After fine-mapping to identify independently significant variants within each locus, we mapped all significant DNA variants to a total of 432 genes, which in aggregate were found to be involved in biological processes related to extracellular matrix maintenance (particularly elastin synthesis and degradation), colonic structure, and colonic motility.

Several analyses taken together support the notion that the genetic variation discovered herein manifests biologically in alterations of colonic structure and function. First, the genes that we proposed are involved in the development of diverticular disease were not only found to be enriched for expression in the sigmoid colon, but also in organs (such as the esophagus and the aorta) that have walls with prominent smooth muscle and extracellular matrix (ECM) components, similar to the colon. Next, the phenotypes with the best genetic correlation to diverticular disease were connective tissue disorders, such as hernias, pelvic organ prolapse, and aortic aneurysms; not inflammatory phenotypes like ulcerative colitis or Crohn’s disease. Third, our single cell RNAseq analysis revealed that the genes we’ve identified as being associated with diverticular disease are normally expressed in the sigmoid colon in the specific cells that are responsible for colonic motility (the interstitial cells of Cajal and smooth muscle cells) and colonic structure (smooth muscle cells and fibroblasts). Finally, we performed proteome-wide Mendelian randomization analysis that linked the genetic variation we associated with diverticular disease to predicted plasma protein levels, finding numerous overlapping protein targets (such as elastin) including some which are the targets of current pharmaceuticals, raising the possibility of drug repurposing. For example, everolimus, a mammalian target of rapamycin (mTOR) antagonist, was shown to rescue defective elastin phenotypes in stem cell-derived vascular smooth muscle cells in individuals with *ELN* gene.^49^ Though the risk profile of everolimus is too unfavorable to consider using it to prevent recurrent diverticulitis, it illustrates the possibility of using drugs targeted toward correcting elastin deficiency, rather than using surgery, to prevent diverticulitis.

Some of the individual genes identified in our analysis that can be viewed as examples of genes which, if altered, may contribute to diverticular disease include: *ANO 1* (which contributes to intestinal pacemaker activity), *ELN* (which is encodes elastin found in the extracellular matrix), *COL15A1* (which encodes collagen and contributes to cell adhesion and extracellular matrix organization), and *LTBP1* (which contributes to extracellular matrix formation and has known clinical manifestations leading to *cutis laxa* (abnormally loose skin), inguinal hernias, and tortuous arteries).

The analysis to this point applies to diverticular disease writ large, rather than to diverticulitis specifically. However, diverticulosis is an asymptomatic and underdiagnosed disease and diverticular disease diagnoses come to light primarily through symptomatic diverticulitis^50^ or through screening colonoscopy (for which only 61% of the eligible population is up-to-date, and which is an imperfect tool to diagnose diverticulosis).^51,52^ Therefore, we hypothesize that our identified DD cases are enriched for symptomatic disease, which is supported by the fact that a 12% DD prevalence in our population is substantially lower than population-level estimates of asymptomatic diverticulosis.^53^ We suggest therefore that the above-described analyses are more likely to reflect genetic variation associated with diverticulitis than with diverticulosis.

Additionally, our immunohistochemical analysis provides evidence that diverticulitis itself is a manifestation of disordered colonic structure, above and beyond any alterations that produce diverticulosis. Specifically, we found dramatically reduced elastin density (adjusted for age and sex) in the sigmoid colon muscularis propria of people requiring emergency surgery for severe diverticulitis not only compared to controls without diverticular disease, but also compared to those with diverticulosis. This supports the possibility that loss of elastin in the muscularis propria increases the risk of diverticulitis directly, rather than increasing the risk of diverticulitis by predisposing to diverticulosis. Importantly, this analysis was conducted when comparing the healthiest part of the colon in diverticulitis patients (the surgical margin, i.e. the area furthest away from the area of diverticulitis) which is least likely to be affected by local inflammation, to the peri-diverticular area in diverticulosis patients. This implies a widespread colonic elastin deficit in diverticulitis patients compared to diverticulosis patients. Equally importantly, none of the diverticulitis patients requiring emergency surgery had documented prior episodes of diverticulitis or gastrointestinal inflammatory disorders, meaning that their elastin deficit was unlikely to be the result of chronic inflammation secondary to disease and more likely to be a (perhaps genetically-mediated) condition that pre-dated their diverticulitis.

In conclusion, it has never been clear whether processes such as inflammation lead to weakening of the colonic wall and the development of DD, or whether connective tissue deficiency is the primary insult leading to DD which then makes a person susceptible to perforation and inflammation. Our analysis suggests that genetic alterations lead to abnormal colonic wall extracellular matrix composition which predisposes to DD, and identifies specific cell types (fibroblasts and smooth muscle cells) and biological processes (extracellular matrix and elastin formation and degradation) involved in this disordered development. This interpretation is further supported by our LDSC results showing shared genetic liability between DD and other connective tissue and smooth muscle diseases (e.g. hernia, pelvic organ prolapse, and abdominal aortic aneurysm).

One of the main limitations of this study is that it relies for the identification of diverticular disease on electronic medical record coding using Phecodes that do not distinguish between diverticulosis and diverticulitis, but as discussed above we believe our data is enriched for diverticulitis rather than diverticulosis. Nevertheless, disentangling the genetic etiology of symptomatic from asymptomatic diverticulosis remains a challenge. A further limitation is that the largest contributor to our meta-analysis was a GWAS in the VA Million Veteran Program, which is overwhelmingly male,^54^ and that the current study does not include significant numbers of individuals of Asian ancestry, whose anatomically right-sided diverticular disease may stem from a different genetic etiology.^55^

## Data Availability

All data produced in the present study will be made available upon publication in a peer-reviewed journal

## Acknowledgements & declarations

The authors would like express their gratitude for the participants of the Penn Medicine Biobank. The PMBB is supported by Perelman School of Medicine at University of Pennsylvania, a gift from the Smilow family, and the National Center for Advancing Translational Sciences of the National Institutes of Health under CTSA award number UL1TR001878.

The authors thank Million Veteran Program (MVP) staff, researchers, and volunteers, who have contributed to MVP, and especially participants who previously served their country in the military and now generously agreed to enroll in the study.^54^ (See https://www.research.va.gov/mvp/ for more details). This research is based on data from the Million Veteran Program, Office of Research and Development, Veterans Health Administration, and was supported by the Veterans Administration (VA) Million Veteran Program (MVP) award #000. This research is based on dbGaP data accession number phs001672.v11.p1.

The current study was approved by the University of Pennsylvania IRB Protocol # 850391. Claude AI (versions 3.5 Sonnet and Sonnet 5) was used as an aid for writing computationally efficient and accurate code in analysis and figure generation; the authors take final responsibility for the integrity of any and all content generated by this tool. All computational code used for analysis will be made publicly available upon publication of this manuscript.

## Abbreviations

DD: Diverticular disease
FUMA: Functional Mapping and Annotation of Genome-Wide Association Studies
GWAS: Genome wide association study
ICD: International classification of diseases
MAF: Mean allele frequency
MAF: Mean allele frequency
MVP: Million veterans program
PIP: Posterior inclusion probability
PMBB: Penn medicine biobank
QTL: quantitiative trait loci
SNP: Single nucleotide polymorphism
UKB: United Kingdom Biobank

## Data Transparency

Data sources are publicly available. Analytic methods will be made available via Github upon acceptance of the manuscript

---

**Extended Data Table 1.** 394 genome-wide significant loci disocvered in the genome-wide association study meta-analysis. (Note: only first page displayed.)

| Chromosome | Position (hg19) | rsid | Effect allele | Other allele | Standard error | p value | Effect (Beta) | Novel? |
| --- | --- | --- | --- | --- | --- | --- | --- | --- |
| 1 | 1299129 | rs2313197 | T | C | 0.0106 | 4.12E-08 | -0.0582 | Yes |
| 1 | 6645867 | rs6682920 | A | G | 0.0043 | 1.73E-08 | -0.0245 | Yes |
| 1 | 10652977 | rs284273 | C | T | 0.0076 | 1.89E-18 | -0.0664 | No |
| 1 | 22379088 | rs2473325 | A | G | 0.0043 | 1.31E-16 | -0.0354 | No |
| 1 | 41276296 | rs114015677 | G | A | 0.0102 | 9.96E-10 | 0.062 | Yes |
| 1 | 59653742 | rs3889199 | G | A | 0.0046 | 2.21E-13 | -0.0337 | Yes |
| 1 | 61824136 | rs7553528 | T | G | 0.0049 | 8.47E-09 | 0.0282 | Yes |
| 1 | 92260920 | rs428815 | T | C | 0.0067 | 5.95E-11 | -0.0441 | Yes |
| 1 | 110364821 | rs2813773 | A | G | 0.0045 | 9.06E-16 | 0.0366 | No |
| 1 | 112832256 | rs4268356 | C | G | 0.0074 | 2.06E-10 | -0.0469 | Yes |
| 1 | 149907259 | rs113759410 | A | G | 0.0093 | 4.38E-08 | -0.0509 | Yes |
| 1 | 150530548 | rs56228576 | G | C | 0.0124 | 1.86E-10 | -0.0789 | Yes |
| 1 | 151258314 | rs116753436 | C | T | 0.0089 | 6.65E-09 | -0.0518 | Yes |
| 1 | 151884499 | rs6672064 | G | A | 0.0046 | 1.90E-29 | -0.0517 | No |
| 1 | 154791995 | rs1218600 | A | C | 0.0045 | 4.40E-08 | -0.0249 | Yes |
| 1 | 156551920 | rs115500783 | G | C | 0.0206 | 8.21E-09 | 0.1189 | Yes |
| 1 | 174172384 | rs2860907 | C | T | 0.0044 | 1.02E-12 | -0.0313 | Yes |
| 1 | 174736877 | rs1653636 | C | T | 0.0044 | 1.65E-11 | 0.0293 | Yes |
| 1 | 179721908 | rs2504047 | A | G | 0.0052 | 5.56E-08 | 0.0285 | Yes |
| 1 | 201539249 | rs987319 | T | G | 0.0082 | 3.80E-09 | 0.0486 | Yes |
| 1 | 203410455 | rs61827971 | T | C | 0.0044 | 2.71E-09 | 0.026 | Yes |
| 1 | 204309134 | rs17405926 | C | T | 0.0046 | 8.07E-15 | 0.036 | Yes |
| 1 | 207536845 | rs11120769 | G | T | 0.0039 | 4.87E-13 | -0.0283 | Yes |
| 1 | 215474906 | rs11120546 | G | A | 0.0041 | 1.03E-08 | -0.0238 | Yes |
| 1 | 219294570 | rs61823192 | T | C | 0.0158 | 1.39E-43 | -0.219 | No |
| 1 | 220023215 | rs61836039 | A | C | 0.0174 | 6.14E-13 | -0.1253 | No |
| 1 | 221071242 | rs2491255 | G | T | 0.004 | 1.22E-21 | -0.0382 | No |
| 1 | 234353053 | rs10910384 | C | A | 0.0049 | 1.43E-112 | 0.1108 | No |
| 1 | 237278525 | rs2485558 | C | G | 0.0048 | 3.93E-18 | -0.0415 | Yes |
| 1 | 237989132 | rs12024181 | T | A | 0.0044 | 1.08E-11 | -0.03 | Yes |
| 1 | 244377774 | rs10737892 | T | G | 0.0042 | 5.46E-09 | 0.0245 | Yes |
| 1 | 245773041 | rs12041565 | T | C | 0.0052 | 1.89E-22 | 0.0511 | No |
| 2 | 2297348 | rs55933406 | G | C | 0.0044 | 3.93E-09 | 0.0258 | Yes |
| 2 | 11317159 | rs72785462 | C | T | 0.0078 | 3.46E-08 | -0.043 | Yes |

**Extended Data Table 2.**
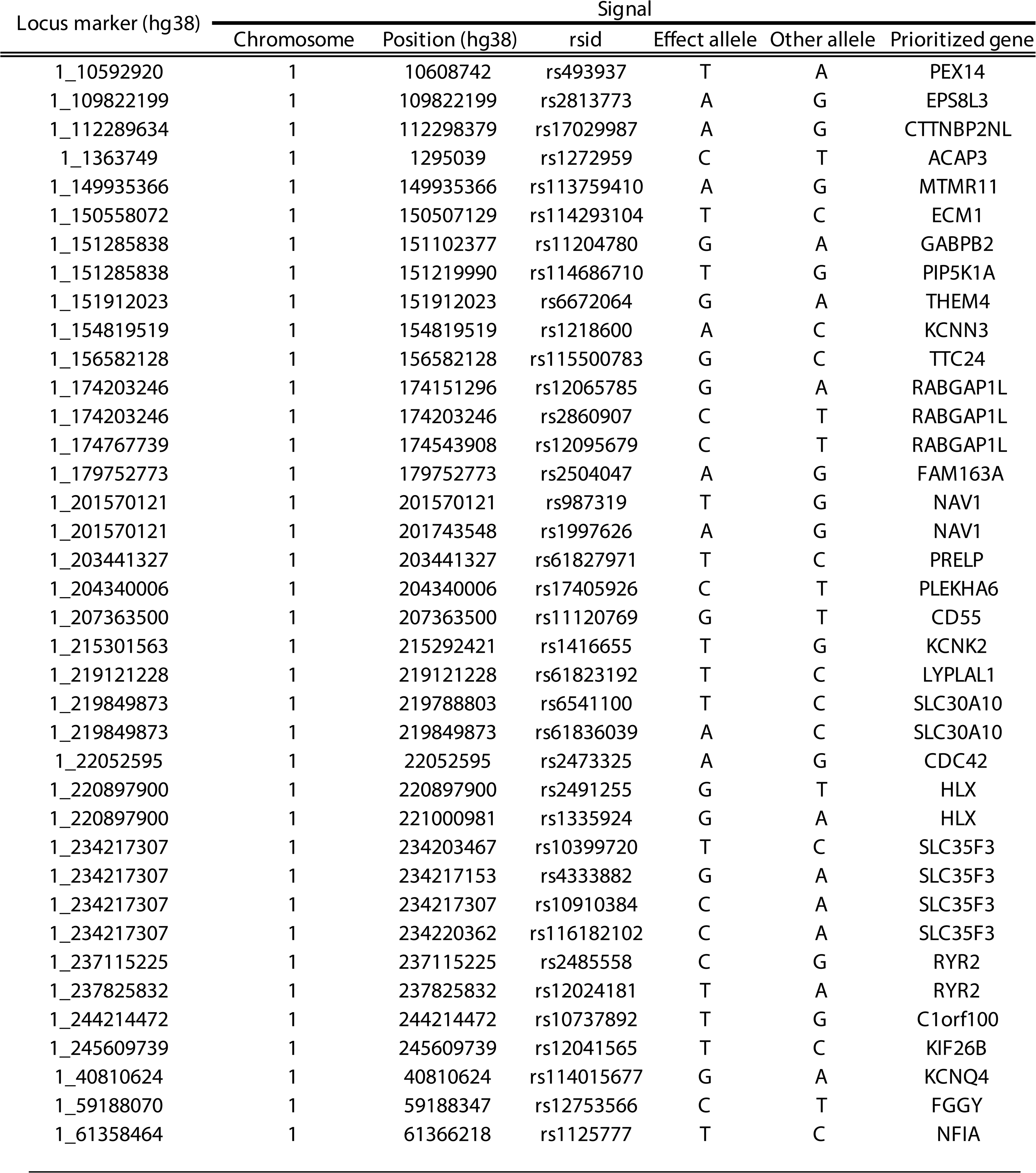
Genes prioritized for each signal. Locus marker denotes the index variant (form atted as chromosome_position) that defines the locus in which the signal resides (Note: only first page displayed)

**Extended Data Fig. 1.**
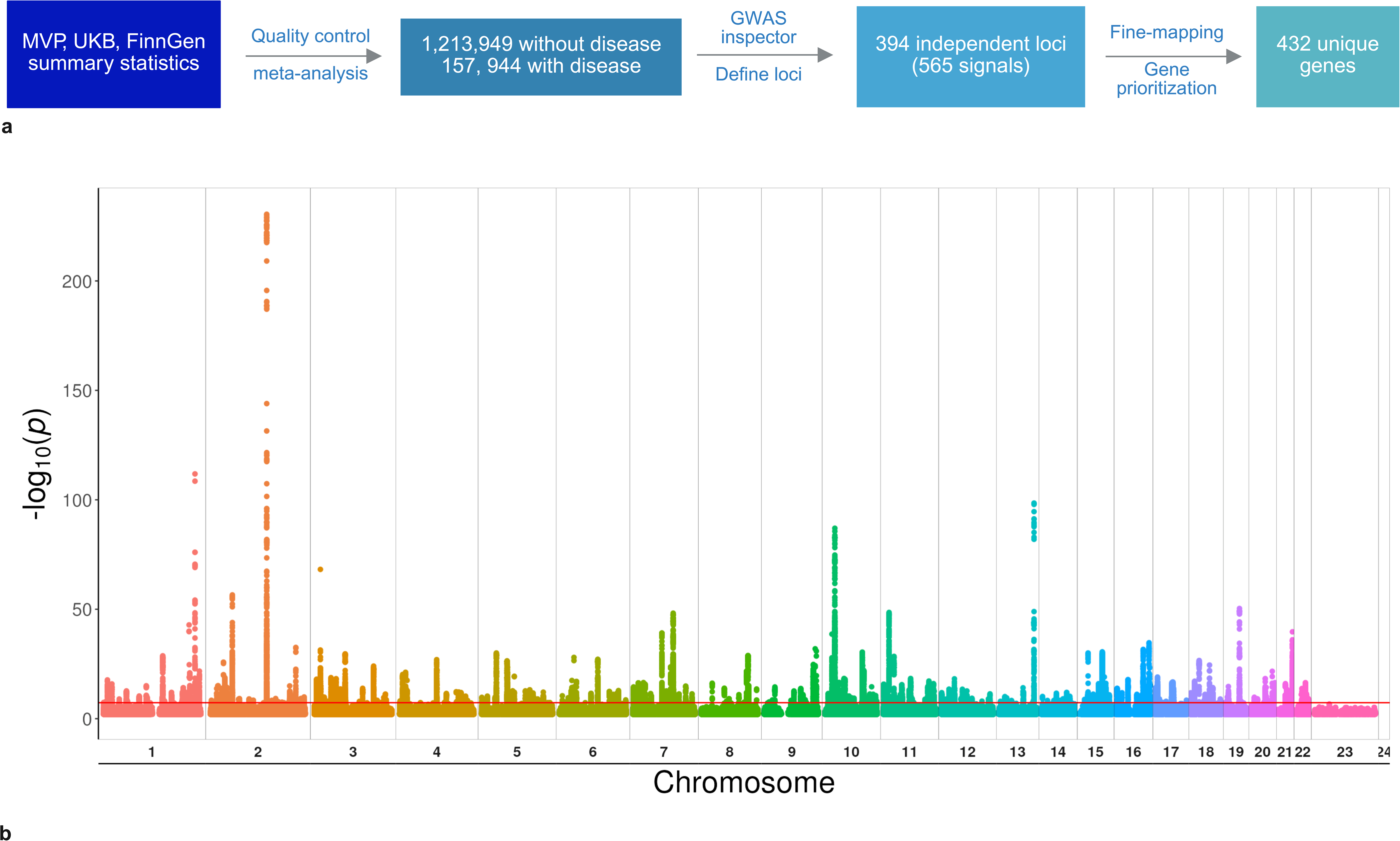
Study overview. **a,** Flowchart of GWAS and gene prioritization. **b,** Manhattan plot demonstrates results of GWAS meta-analysis.

**Extended Data Fig. 2.**
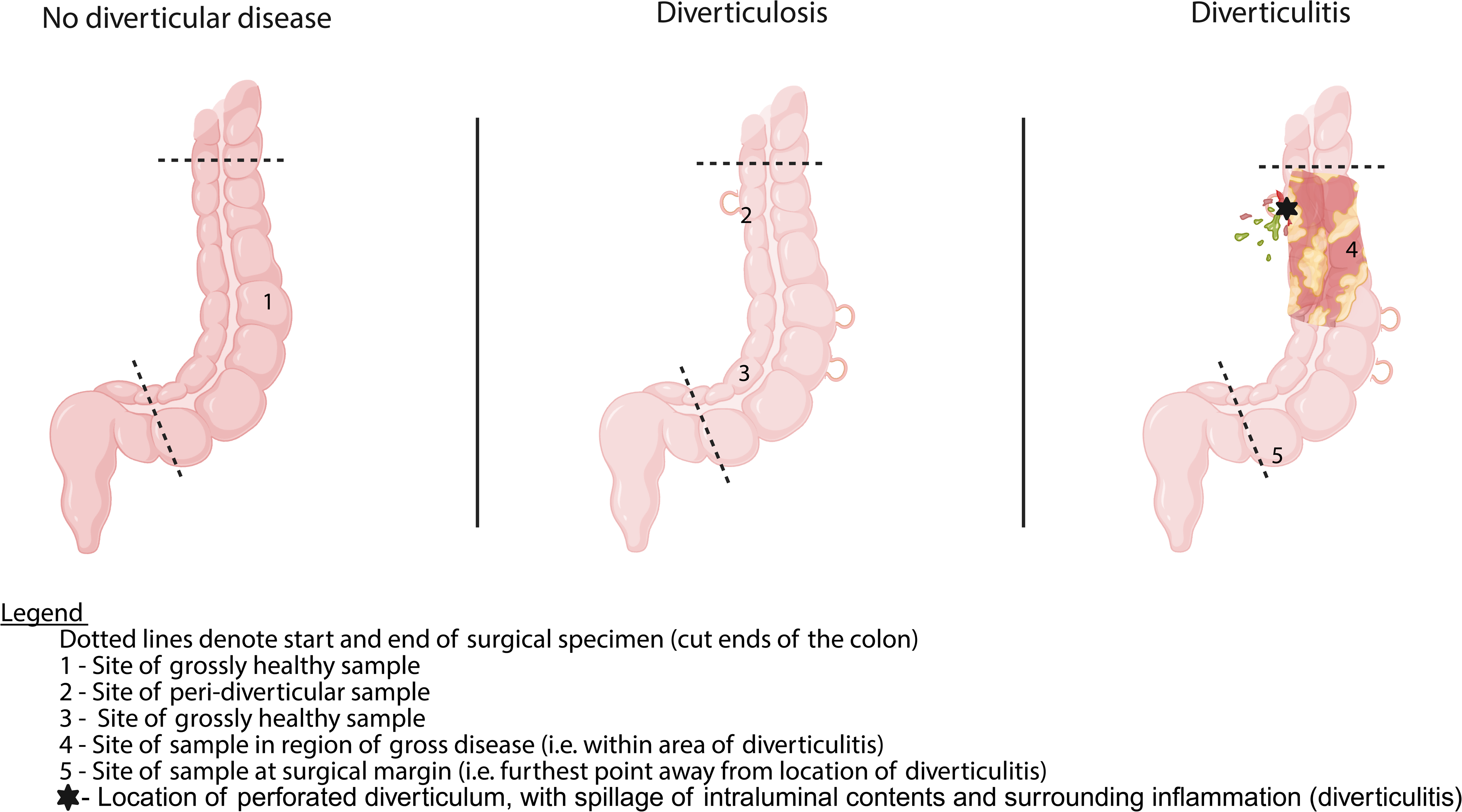
Locations of samples. taken from surgical specimens for immunohistochemistry staining and elastin quantification. Dotted lines denote start and end of surgical specimen (cut ends of the colon) 1 - Site of grossly healthy sample 2 - Site of peri-diverticular sample 3 - Site of grossly healthy sample 4 - Site of sample in region of gross disease (i.e. within area of diverticulitis) 5 - Site of sample at surgical margin (i.e. furthest point away from location of diverticulitis) ★ - Location of perforated diverticulum, with spillage of intraluminal contents and surrounding inflammation (diverticulitis)

**Extended Data Fig. 3.**
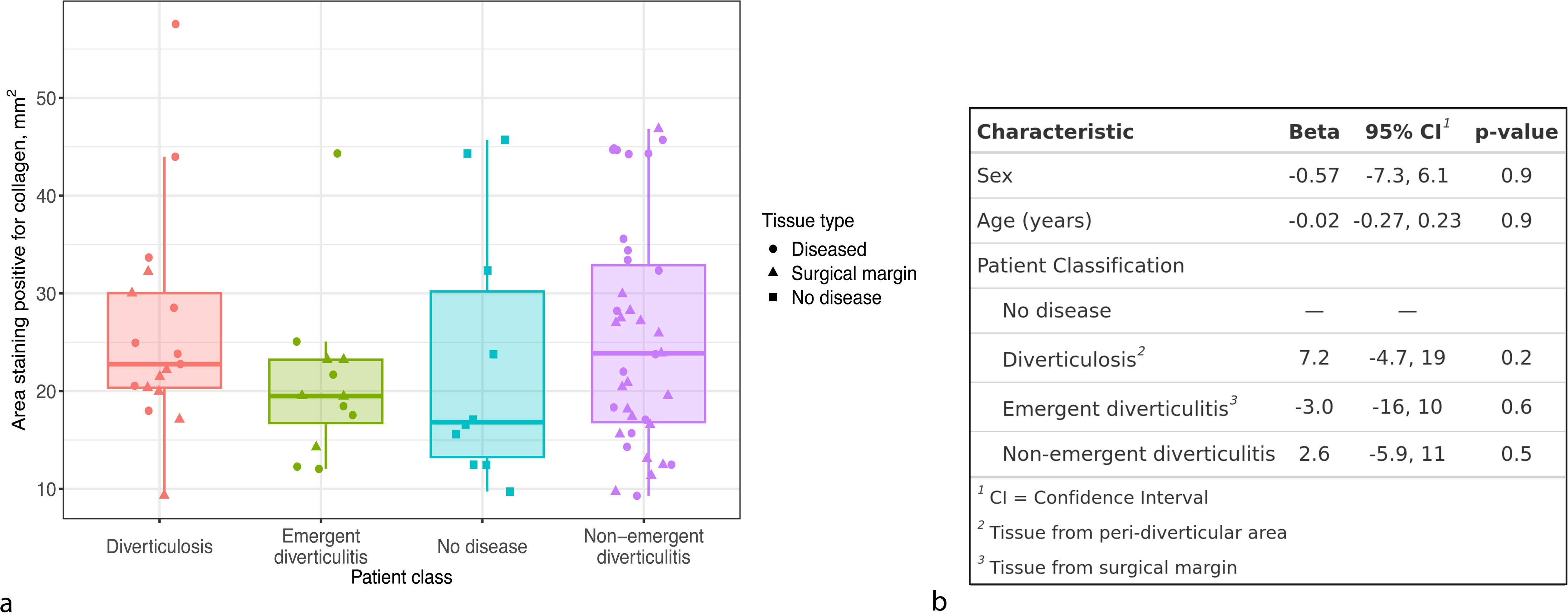
Collagen quantification. **a,** Each dot represents the measured collagen density on a prepared slide, with boxplot showing median collagen density (horizontal line) by patient class, interquartile range (IQR; the borders of the box) and whiskers extending 1.5*IQR from the end of the box. **b,** Multivariable linear regression showing no significant difference in collagen density among patients undergoing emergency diverticulitis surgery compared to those without disease. Peri-diverticular area corresponds to location 2 in Extended Data Fig.6; surgical margin corresponds to location 5 in Extended Data Fig.6.

**Extended Data Fig. 4.**
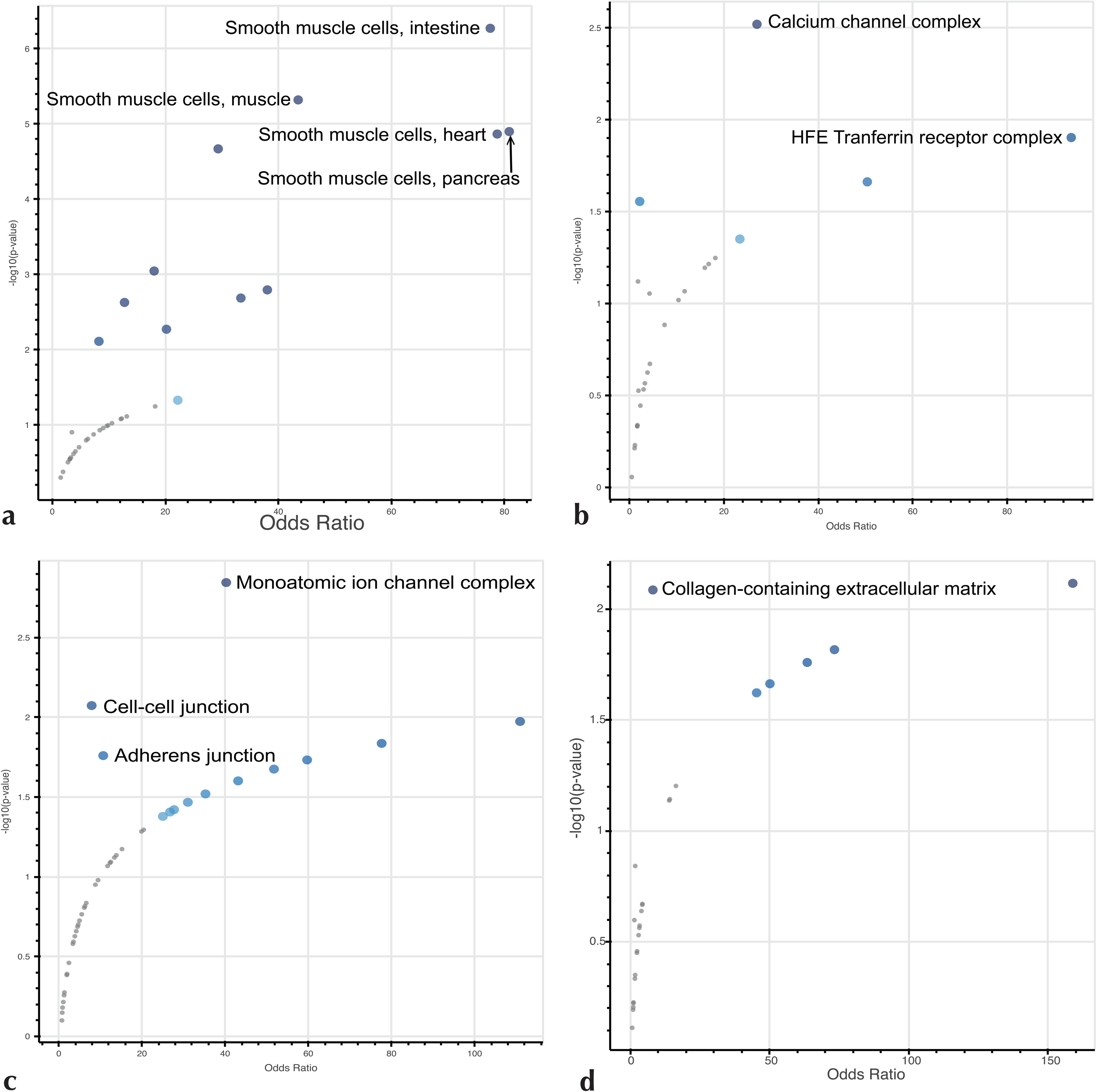
Pathway analysis of clusters of genes. enriched in **a,** smooth muscle cells (cluster 11, Wright et al.), in Descartes Cell Types enrichr pathway, **b,** interstitial cells of Cajal (ICC) (cluster 10, Wright et al.), in Gene Ontolocy (GO) cellular component 2025, **c,** ICC (cluster 6, Hickey et al.), in GO cellular component 2025, **d,** WNT5B+ fibroblasts (cluster 10, Hickey et al.), in GO cellular component 2025

**Extended Data Fig. 5.**
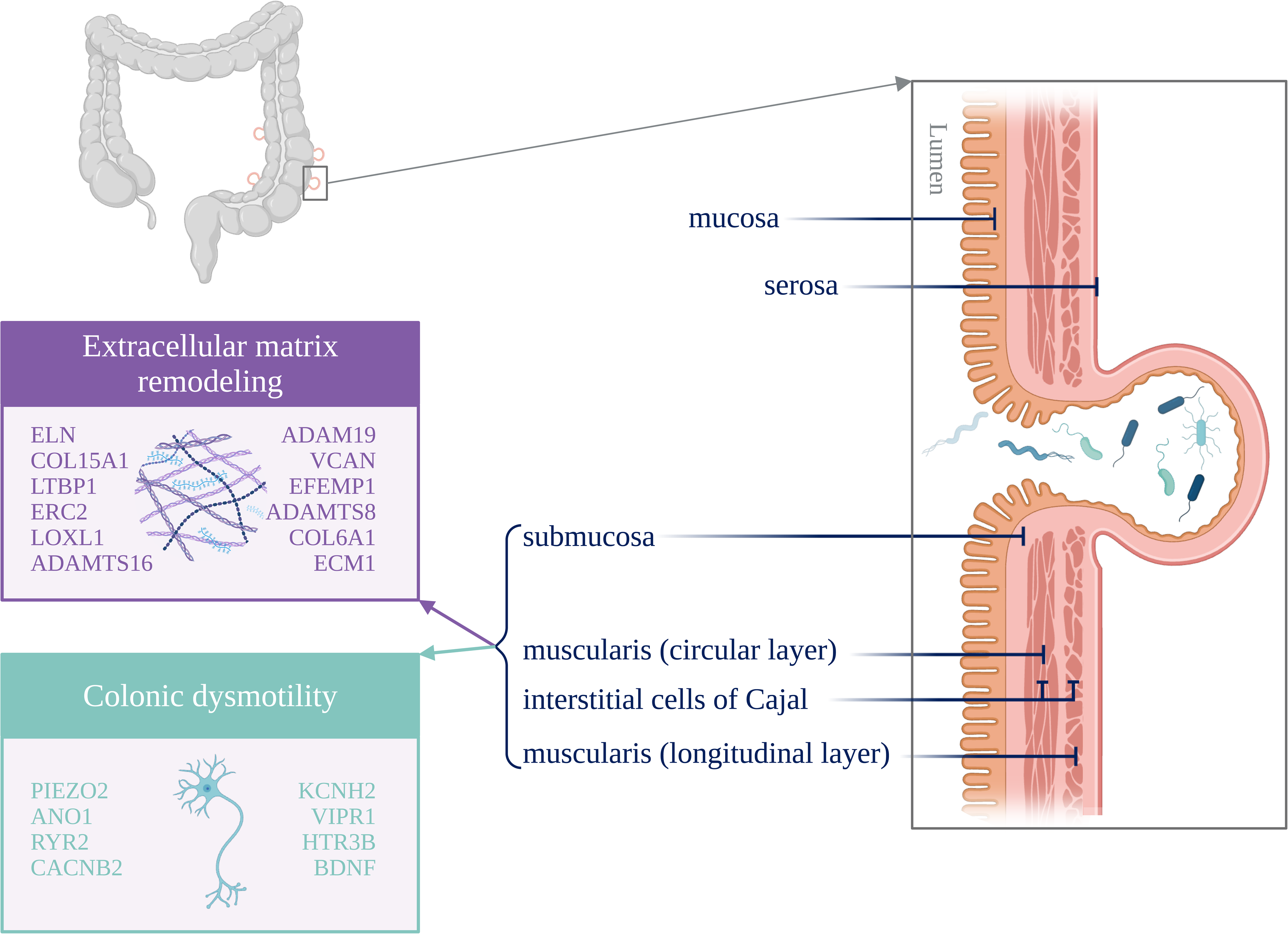
Proposed biological changes through which the genes identified in this GWAS may increase liability to diverticular disease.

**Supplementary Data Table 1.**
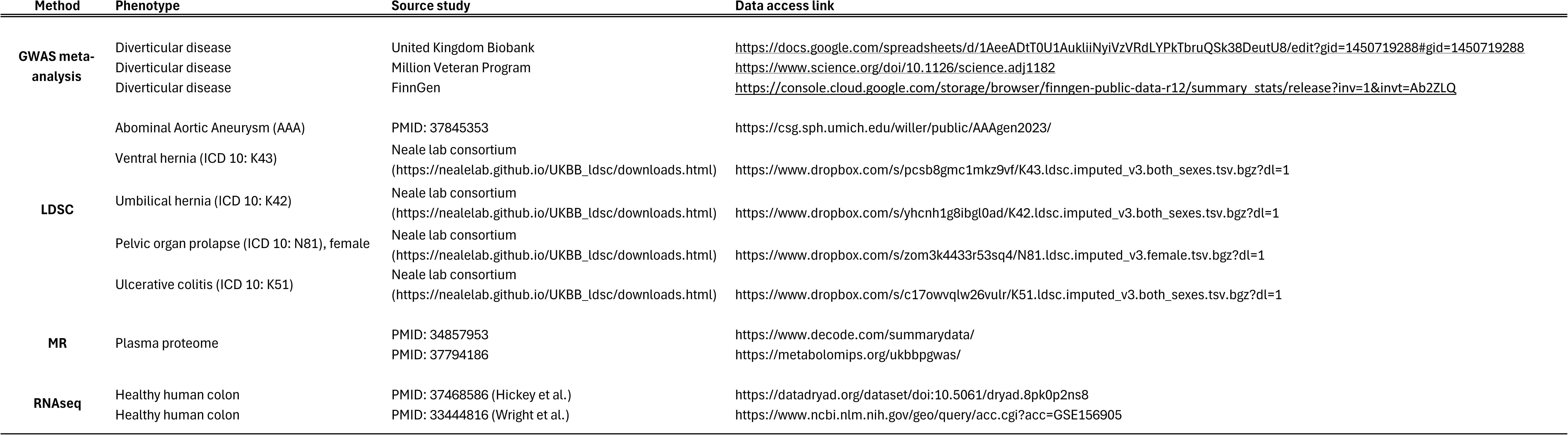
Input data sources. LDSC = Linkage disequlibrium score regression, MR = Mendelian randomization, PMID = PubMed identifier

**Supplementary Data Table 2.**
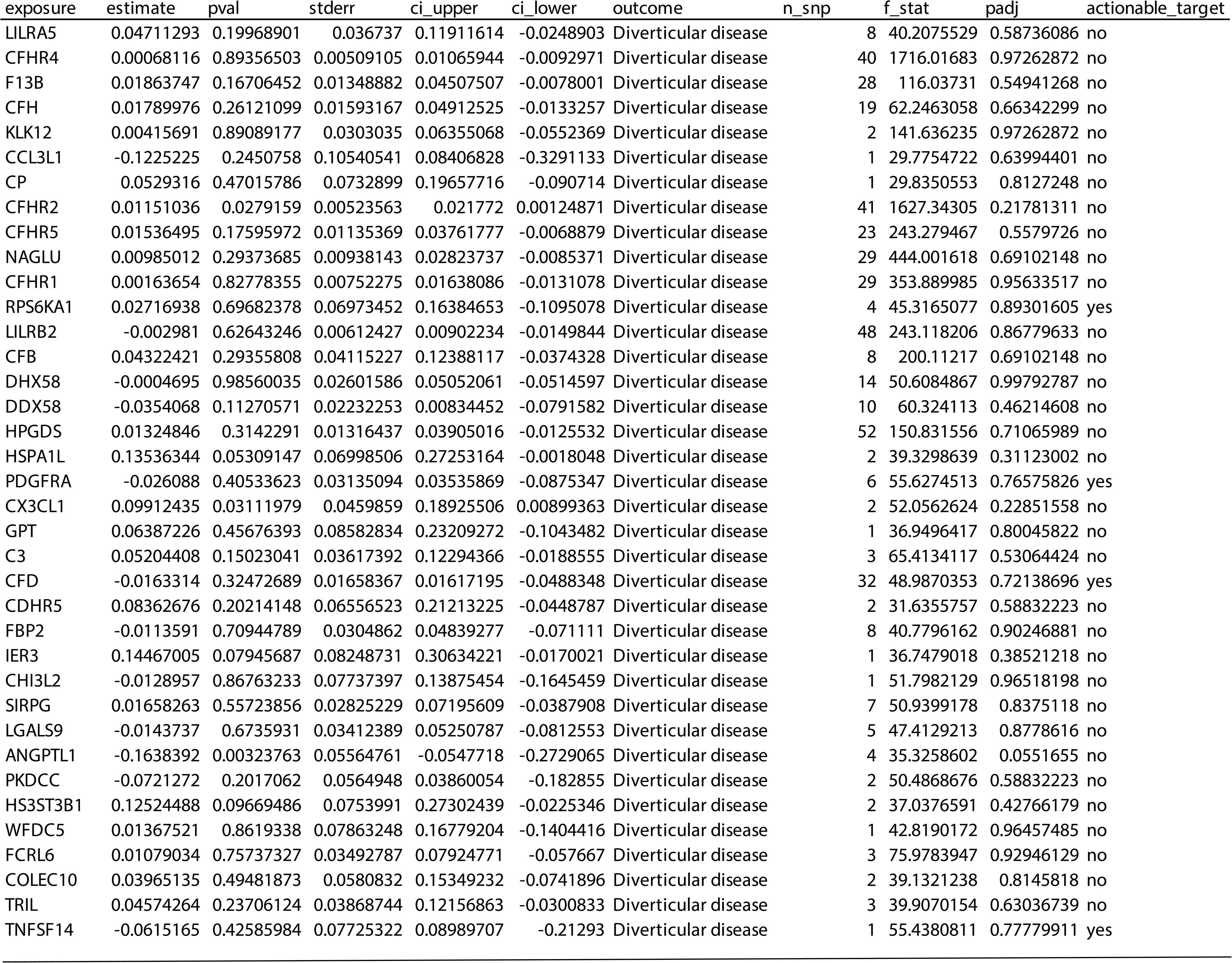
Full results of pQTL MR using deCODE data. (Note: only first page displayed.)

**Supplementary Data Table 3.** Full results of pQTL MR using UK Biobank Pharma Proteomics Project (UKB-PPP) data. (Note: only first page displayed.)

**Supplementary Data Table 2.** Full results of pQTL MR using deCODE data. (Note: only first page displayed.)
| exposure | estimate | pval | stderr | ci_upper | ci_lower | outcome | n_snp | f_stat | padj | actionable_target |
| --- | --- | --- | --- | --- | --- | --- | --- | --- | --- | --- |
| NTRK2 | 0.07441101 | 0.13813318 | 0.05018348 | 0.17276882 | -0.0239468 | Diveticular disease | 7 | 28.988781 | 0.49616266 | yes |
| NTF3 | 0.01953232 | 0.35875327 | 0.02128298 | 0.0612462 | -0.0221816 | Diveticular disease | 16 | 59.0297798 | 0.72920389 | no |
| APBB1IP | -0.0040055 | 0.87164682 | 0.02479137 | 0.04458472 | -0.0525957 | Diveticular disease | 11 | 92.7752352 | 0.96755322 | no |
| FSHB | 0.02396781 | 0.68618828 | 0.05932139 | 0.1402356 | -0.0923 | Diveticular disease | 9 | 50.710388 | 0.89322283 | no |
| CDH2 | 0.05151839 | 0.36719772 | 0.05713253 | 0.16349609 | -0.0604593 | Diveticular disease | 7 | 65.1202918 | 0.73270344 | no |
| DYNLT1 | 0.03713394 | 0.58109568 | 0.06729784 | 0.16903528 | -0.0947674 | Diveticular disease | 2 | 44.5262171 | 0.84636014 | no |
| PSG1 | 0.00035019 | 0.95384572 | 0.00605045 | 0.01220886 | -0.0115085 | Diveticular disease | 82 | 267.651328 | 0.98991844 | no |
| BAG3 | 0.10185327 | 0.05209106 | 0.05243734 | 0.20462857 | -0.000922 | Diveticular disease | 3 | 39.4015579 | 0.3200335 | no |
| NRP2 | 0.02488735 | 0.20886678 | 0.01980392 | 0.06370233 | -0.0139276 | Diveticular disease | 24 | 62.5192435 | 0.5835809 | no |
| VEGFC | 0.02483962 | 0.21830393 | 0.02017758 | 0.06438695 | -0.0147077 | Diveticular disease | 2 | 133.832634 | 0.58870142 | yes |
| SERPINE2 | 0.00501667 | 0.63823058 | 0.01066981 | 0.02592912 | -0.0158958 | Diveticular disease | 50 | 93.1940517 | 0.87120024 | no |
| SERPINE1 | 0.02149211 | 0.57675436 | 0.03850723 | 0.09696489 | -0.0539807 | Diveticular disease | 7 | 45.9628951 | 0.84375773 | yes |
| SDC4 | -0.0003172 | 0.98442985 | 0.01625228 | 0.03153671 | -0.0321711 | Diveticular disease | 20 | 60.9846111 | 0.99477162 | no |
| PPBP | 0.0134705 | 0.73194347 | 0.03932517 | 0.09054642 | -0.0636054 | Diveticular disease | 1 | 48.3906988 | 0.91819694 | no |
| PDGFA | -0.1096041 | 0.32224856 | 0.11072828 | 0.1074193 | -0.3266276 | Diveticular disease | 1 | 35.0920918 | 0.6995454 | no |
| DKK1 | -0.0348251 | 0.20561285 | 0.027514 | 0.01910139 | -0.0887515 | Diveticular disease | 7 | 58.1549923 | 0.58109264 | yes |
| CCN2 | 0.02544539 | 0.14732002 | 0.01755989 | 0.05986215 | -0.0089714 | Diveticular disease | 25 | 86.4769037 | 0.51379162 | no |
| BDNF | -0.0039665 | 0.95469003 | 0.06981024 | 0.13285907 | -0.1407921 | Diveticular disease | 1 | 43.0092164 | 0.98991844 | no |
| APP | 0.04598154 | 0.2333083 | 0.038579 | 0.121595 | -0.0296319 | Diveticular disease | 4 | 43.6897602 | 0.60703364 | yes |
| ANGPT1 | 0.01814175 | 0.75856986 | 0.05902457 | 0.13382778 | -0.0975443 | Diveticular disease | 3 | 31.1978204 | 0.92669805 | yes |
| SEMA4D | 0.03231407 | 0.45664594 | 0.04341066 | 0.1173974 | -0.0527693 | Diveticular disease | 7 | 46.2580803 | 0.79114879 | no |
| CBLN4 | 0.00955797 | 0.56141732 | 0.01645833 | 0.0418157 | -0.0226998 | Diveticular disease | 35 | 60.8787101 | 0.83533122 | no |
| B4GAT1 | -0.0076451 | 0.79111298 | 0.02886397 | 0.04892726 | -0.0642174 | Diveticular disease | 24 | 64.5785114 | 0.93573015 | no |
| GUSB | -0.0422998 | 0.39493098 | 0.04972297 | 0.05515544 | -0.139755 | Diveticular disease | 6 | 75.5430022 | 0.75194204 | no |
| DCBLD2 | 0.02123831 | 0.19814844 | 0.01650418 | 0.05358591 | -0.0111093 | Diveticular disease | 21 | 112.616277 | 0.57163948 | no |
| KLK7 | 0.01905582 | 0.13668688 | 0.01280424 | 0.04415167 | -0.00604 | Diveticular disease | 17 | 210.68585 | 0.49616266 | no |
| TNFSF12 | 0.07570017 | 7.38E-09 | 0.01309246 | 0.10136092 | 0.05003943 | Diveticular disease | 38 | 81.9463437 | 1.82E-06 | yes |
| IL1R1 | 0.00062237 | 0.97074825 | 0.01697227 | 0.0338874 | -0.0326427 | Diveticular disease | 27 | 45.2913275 | 0.99166255 | yes |
| FGFR4 | 0.01062476 | 0.02535046 | 0.00475164 | 0.01993781 | 0.00131172 | Diveticular disease | 92 | 224.621296 | 0.20561785 | yes |
| NAGK | -0.0332018 | 0.17866221 | 0.02468748 | 0.01518474 | -0.0815884 | Diveticular disease | 17 | 63.9116912 | 0.55247322 | no |
| OGN | -0.0226444 | 0.01038278 | 0.00883576 | -0.0053266 | -0.0399622 | Diveticular disease | 53 | 126.654467 | 0.11593196 | no |
| FSTL1 | -0.0145695 | 0.52261988 | 0.02278938 | 0.03009684 | -0.0592359 | Diveticular disease | 10 | 62.3906516 | 0.82054643 | no |
| FNDC1 | 0.00936593 | 0.40979983 | 0.01136302 | 0.03163704 | -0.0129052 | Diveticular disease | 50 | 103.123071 | 0.76188682 | no |
| FAP | 0.02132292 | 0.12336337 | 0.01383878 | 0.04844644 | -0.0058006 | Diveticular disease | 10 | 108.822312 | 0.48121397 | yes |
| CCN1 | -0.037161 | 0.22863937 | 0.03086789 | 0.02333899 | -0.0976609 | Diveticular disease | 13 | 40.3569624 | 0.60278479 | no |

